# High-Resolution District Level Contraceptive Prevalence in Pakistan Using a Bayesian Small Area Estimation Approach

**DOI:** 10.64898/2026.02.25.26347119

**Authors:** Muhammad Ibrahim, Olan Naz, Ayesha Javeed, Aisha Irum, Ayesha Khan, Adnan Ahmad Khan

## Abstract

**Introduction:** National surveys in Pakistan are typically representative only at national or provincial levels, leaving large uncertainties in district-level contraceptive prevalence. This obscures local heterogeneity and limits data-driven program planning. Administrative data, although more frequent and detailed, are often underused due to reporting and measurement challenges. This study develops a multi-source small area estimation (SAE) framework to generate district-level estimates of contraceptive prevalence rate (CPR) and modern contraceptive prevalence rate (mCPR) using routine commodities data.

**Methods:** A two-stage Bayesian SAE model was constructed to integrate survey, supply, and census data. In Stage 1, contraceptive dispensation data from the Contraceptive Logistics Management Information System (cLMIS) were converted into inferred users, normalized to married women of reproductive age (MWRA) from the 2023 Census, and scaled to provincial CPR benchmarks from the Pakistan Social and Living Standards Measurement Survey (PSLM). In Stage 2, a bivariate hierarchical Bayesian model jointly estimated CPR and mCPR, accounting for measurement error and borrowing statistical strength from socioeconomic and demographic covariates. Convergence and model stability were assessed through standard diagnostics (R-hat, ESS, BFMI, divergence checks).

**Results:** District-level estimates were produced for 121 districts. CPR ranged from 9% to 46% and mCPR from 6% to 35%. Aggregated provincial estimates were consistent with PSLM benchmarks (within ± 0.6 percentage points). Comparison with published district studies showed mean absolute deviations around 4 percentage points.

**Conclusion:** The Bayesian SAE framework generates statistically coherent, high-resolution contraceptive prevalence estimates, substantially improving visibility into geographic inequities in Pakistan’s family planning landscape. These granular metrics offer policymakers an actionable basis for prioritizing underserved districts and tailoring context-sensitive interventions.

## Introduction

Reliable and spatially detailed health data are essential for equitable and effective public health planning. For reproductive health indicators such as the Contraceptive Prevalence Rate (CPR), national or provincial survey estimates often mask considerable variation across districts. In Pakistan, as in many low- and middle-income countries (LMIC), health outcomes and resource allocation vary widely by geography, socioeconomic status, and rural–urban context, underscoring the need for subnational, data-driven decision-making (1, 2).

Although Pakistan routinely collects extensive service and supply data, these administrative records are seldom analyzed for policy because of concerns about completeness, quality, and usability. Weak linkage between administrative data and population-based sources such as censuses and national surveys further limits their reliability (3, 4). Instead, health indicators are largely reported through surveys such as the Pakistan Demographic and Health (PDHS) or Pakistan Social and Living Standards Measurement (PSLM) Surveys, both of which report only at national or provincial levels. The district-level version of the PSLM does not include family planning and other indicators limiting the ability to make district level inferences. Ad hoc, project-based surveys provide some local information (5, 6), that is not administrative area representative and one point in time snapshots of indicators and service utilization in a narrow geographic coverage and are methodologically heterogeneous (7–11).

As a result, family-planning programs often depend on outdated survey estimates or administrative ratios that do not reflect local operational realities (12). Small area estimation (SAE) methods can address these limitations by statistically combining survey and administrative data to “borrow strength” across areas, producing coherent subnational estimates (13–17). The core strength of this approach lies in the strategic integration of multiple data sources, in our case survey, census, and programming data. In addition, SAE can allow incorporation of data correction protocols allowing such “noisy” data to be converted into reliable, detailed estimates without costly surveys. The use of multiple data source models overcomes issues of accuracy on single source surveys that can make them contentious, by correcting biases more effectively and better reflect the true contraceptive prevalence or other indicator values (3).

These techniques have been applied successfully to diverse health indicators, child mortality, immunization, and poverty, demonstrating their capacity to enhance geographic resolution without new large-scale surveys (18–20). Previous SAE examples have largely focused on smoothing survey gaps (21, 22). In low-data environments such as Pakistan, SAE offers an efficient and replicable means of producing granular reproductive health statistics that can inform Primary Health Care (PHC) planning and monitoring at the operational level.

This study applies and extends SAE to reproductive-health monitoring in Pakistan through a two-stage Bayesian framework consistent with the Fay–Herriot area-level model (21). The framework treats contraceptive logistics data (cLMIS) as a primary district-level signal, explicitly models its measurement error, and jointly estimates CPR and modern CPR (mCPR). By integrating cLMIS, census, and PSLM data with socioeconomic covariates, it generates coherent, policy-relevant estimates for all districts. This methodological contribution demonstrates how routinely collected administrative data can be leveraged to produce high-resolution reproductive-health statistics, advancing Pakistan’s progress toward Sustainable Development Goal 3: Good Health and Well-Being.

## Methodology

### Data Sources in Pakistan

Pakistan has conducted multiple national surveys and maintains over a hundred program datasets covering various health domains. Among these, the Demographic and Health Survey (DHS) and the Pakistan Social and Living Standards Measurement Survey (PSLM)—part of the World Bank’s Living Standards Measurement Study—collect information on contraceptive use. Both surveys employ similar sampling frames and are representative at national and provincial levels, yielding broadly comparable results for family planning and immunization indicators (3). The DHS is conducted every 5-6 years. PSLM is conducted by the Pakistan Bureau of Statistics and the national/provincial survey alternates with a district survey, usually every other year. The national survey has more in depth questionnaire from around 25,000 respondents while the district survey has fewer indicators, including no family planning ones, but is administered to around 180,000 households.

**Figure 1:**
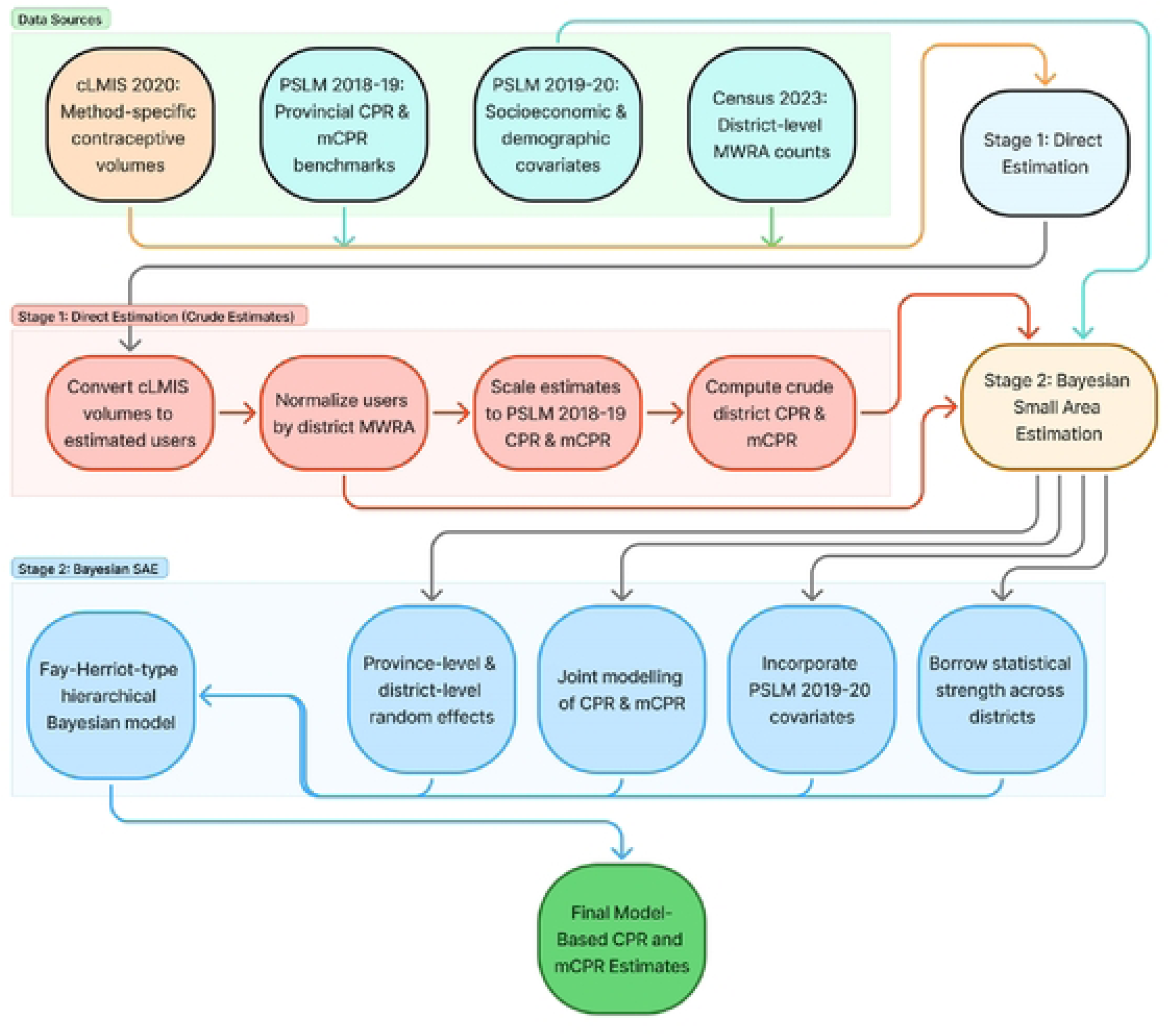
Logical Framework of the District-Level CPR Estimation Process.

Among program datasets, the Contraceptive Logistics Management System (cLMIS) tracks national FP commodities supplies from a central warehouse to each district, and facility. For most part all of national FP commodities are channeled through this system, although private sector supplies are less well documented. Additionally, it does not provide actual users, method continuation and there are other data errors and logistical reporting errors that induce uncertainty require corrective imputations, adjustment, and validation.

### Data Sources Used

We used cLMIS to estimate district prevalence of CPR and mCPR. Data used were from 2020, since that is the last year when a provincial PSLM is available that could be used to validate the model. This study utilized four complementary data sources: 1) the PSLM/HIES (2018-19) for provincial CPR benchmarks, 2) the cLMIS (2020) for district-level contraceptive dispensation data to incorporate the variation between district patterns for contraceptives, 3) the district level PSLM (2019-20) for the covariates and 4) the 2023 Population and Housing Census for demographic denominators, particularly the number of Married Women of Reproductive Age (MWRA). Together, these datasets provide behavioral, supply-side, and demographic dimensions essential for estimating CPR at the district level. Table 1 shows the sample size and average CPR, mCPR for all provinces.

**Table 1:** Sample Districts and Average CPR, mCPR Across provinces.

| Province | Average CPR (%) | Average mCPR (%) | cLMIS Districts | PSLM Districts |
| --- | --- | --- | --- | --- |
| Punjab | 37.8 | 25.4 | 17 | 36 |
| Sindh | 28.1 | 24.1 | 28 | 28 |
| KP | 29.4 | 18.1 | 28 | 30 |
| Balochistan | 13.6 | 0.99 | 21 | 27 |

### Analysis

Two baseline models were used to guide the development of the final hierarchical Bayesian estimator. First, a direct administrative proxy model converted cLMIS contraceptive dispensation into estimated users, normalized these by district-level MWRA counts, and scaled them to PSLM provincial CPR benchmarks, producing the Stage 1 crude district estimates commonly used as direct estimators in SAE.

Second, these crude estimates were fed into an area-level Fay-Herriot-type model, which combines direct estimates with auxiliary covariates and province-level random effects. This informed the conceptual basis for the final bivariate hierarchical Bayesian model, which extends the Fay-Herriot structure by jointly modelling CPR and mCPR, incorporating measurement errors in cLMIS data, and borrowing strength across provinces and districts.

### Stage 1: Crude District Estimates

The first stage involved estimating crude district-level CPR and mCPR by using cLMIS contraceptive dispensation data and provincial benchmarks from PSLM 2018-19.

### 1.1 District-Level Aggregation

The number of contraceptive users for each district was directly calculated from the dispensation data. Data was available at month level from each district which was aggregated at annual level to estimate yearly CPR and mCPR. Each method was standardized and categorized based on its commodity type (condoms, pills, injections, implants and IUDs) for extract the users using conversion factor (23). The total number of users in each district was then calculated by aggregating method-specific users.

### 1.2 Adjusting for Population Differences

To account for variations in population sizes across districts, we normalize contraceptive users by district population using MWRA estimates from the 2023 Census.

### 1.3 Provincial Benchmarks

The provincial average for contraceptive prevalence is established by computing the mean contraceptive users per MWRA across all districts within a province.

### 1.4 Measuring Deviation from Provincial Average

To quantify district-specific deviations from provincial contraceptive trends, the difference between each district’s users per MWRA and the provincial average users per MWRA was calculated. To account for scale differences, a proportional deviation metric was introduced. To prevent outliers from distorting CPR estimates, extreme deviations were capped. Any proportional deviation greater than 100% (i.e., *R*_*d*_≥1) was excluded from further calculations. The provincial-level CPR estimates are as follows: Khyber Pakhtunkhwa (KP) at 30.6%, Punjab at 38.6%, Sindh at 29.5%, and Balochistan at 13.7%. These benchmarks reflect the overall contraceptive use within each province, capturing both modern and traditional methods as reported in representative household surveys. These provincial CPR values were used to establish district-specific adjustments, ensuring that each district’s CPR estimate is scaled appropriately relative to its province.

### 1.5 Estimating CPR Based on District Deviations

To compute the district-specific CPR adjustment, the provincial CPR was scaled using the adjusted proportional deviation. For administratively merged districts (e.g., Karachi districts), MWRA populations and contraceptive users were summed to ensure alignment with population and contraceptive consumption distributions.

### 1.6 Uncertainty Quantification for Crude Estimates

District-level standard errors were constructed by combining provincial sampling uncertainty with within-province variability. Under the assumption of independence between these sources, standard errors were calculated as:

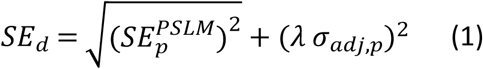

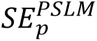 is the design-based standard error from PSLM; *σ*_*adj*,*p*_is the standard deviation of district adjustments within province p; and *λ*, are scaling factors accounting for partial pooling across districts, for CPR was set at 0.75 and for mCPR it was set at 0.80. CPR includes traditional and privately obtained methods, making administrative consumption data noisier; therefore, a slightly stronger pooling factor (0.75) was used. In contrast, modern methods are more consistently recorded through cLMIS, warranting a higher factor (0.80). The higher scaling factor for modern CPR reflects greater within-province heterogeneity in modern method adoption.

To prevent this numerical instability, we imposed minimum standard error thresholds of 1.5 percentage points for CPR and 2.0 percentage points for mCPR. These lower bounds ensure that the model preserves appropriate uncertainty even in data-rich provinces, keeps the district estimates from being over-constrained, and avoids the known issue of underestimating measurement error in administrative-data–anchored small-area estimation models.

### 1.7 Uncertainty in Stage 1 Crude Estimates

Stage 1 estimates are subject to multiple sources of statistical uncertainty that are explicitly addressed in Stage 2. These include measurement error in cLMIS administrative data arising from incomplete private-sector coverage and heterogeneous reporting quality across districts; instability due to the absence of strength borrowing in data-sparse districts; independent estimation of overall and modern CPR despite their underlying linkage; underestimation of uncertainty when administrative measurement error is not incorporated; and the absence of reliable estimates for districts with extreme deviations. The Stage 2 hierarchical Bayesian framework is designed to jointly address these sources of uncertainty through partial pooling, measurement-error modeling, and joint estimation.

### Stage 2: Hierarchical Bayesian Small Area Estimation

We developed a bivariate hierarchical Bayesian model simultaneously estimating overall CPR and modern CPR by integrating four data sources: Stage 1 crude estimates with standard errors, cLMIS administrative data with measurement error modeling, predictor variables from PSLM 2019-20 and PSLM provincial benchmarks with design-based standard errors.

### 2.1 Model Structure

Contraceptive prevalence was modeled as a function of covariates and provincial random effects:

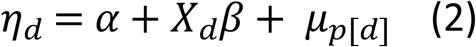

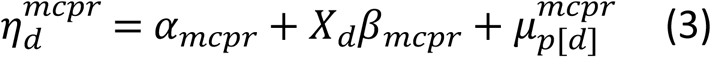

where *η*_*d*_are logit-scale linear predictors, *⍺* are intercepts, *X*_*d*_contains standardized covariates (education index, income index, fertility rate under 25 years old, agriculture proportion and, manufacturing proportion); *β* are the covariate effects; and *μ*_*p*[*d*]_are province-specific random effects. Prevalence rates were obtained via:

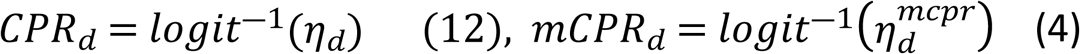

Provincial effects were modeled jointly with constrained correlation:

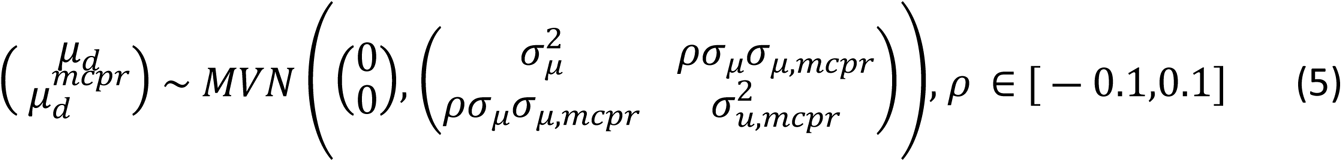

where *σ*_*μ*_ and *σ*_*μ*,*mcpr*_ quantify between-province variability, and *ρ* is constrained to ensure near-independence, allowing overall and modern CPR to exhibit distinct provincial patterns.

cLMIS administrative data were linked to true CPR through dual proxy models with estimated elasticity and heteroskedastic errors:

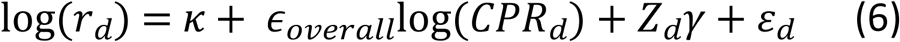

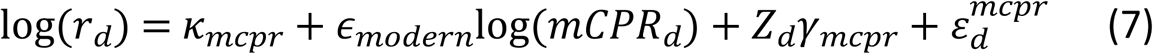

where *r*_*d*_ is observed users per MWRA; *ε* are the elasticity parameters (allowing deviation from unit elasticity); *Z*_*d*_ contains sectoral composition (agriculture, manufacturing); *γ* capture how sectors affect proxy accuracy; and errors follow:

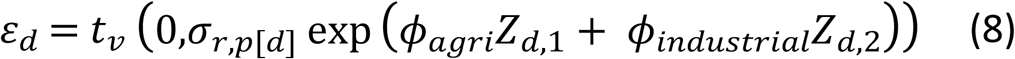

with *σ*_*r*,*p*[*d*]_ representing province-specific baseline error, *ϕ*_*agri*_capturing higher error in agricultural districts, *ϕ*_*industrial*_ capturing lower error in industrial districts, and t-distribution (*v* > 2) providing robustness to outliers.

### 2.2 Data Integration

The model integrates complementary information from three sources through distinct likelihood components. Direct survey-based estimates inform district-level CPR and mCPR where sampling uncertainty is explicitly accounted for. Administrative proxy data from cLMIS provide continuous district-level signals on contraceptive use, with uncertainty modeled to reflect reporting noise. Finally, provincial anchoring constraints ensure that MWRA-weighted district estimates remain coherent with officially reported PSLM provincial CPR and mCPR levels, preserving national and provincial consistency.

### 2.3 Soft Constraint, Covariates Effects and Prior Specification

Modern CPR was softly centered on province-specific modern-to-overall CPR ratios from PSLM, allowing districts to deviate while preserving realistic relationships between overall and modern contraceptive use. A moderate dispersion reflects observed provincial variability, and an upper cap prevents modern CPR from exceeding demographically plausible levels relative to overall CPR.

Contraceptive use is shaped by well-established patterns: education, income, and manufacturing or urban employment are all positively associated with higher CPR, as shown in prior studies (24–34). In contrast, agricultural settings consistently show lower CPR (35–37), while higher fertility, especially among adolescents, also correlates with reduced contraceptive uptake (38–41). These findings justify the direction of priors in our model. Consistent with PSLM data and literature, modern CPR typically represents 60–80% of overall CPR in Pakistan, so priors for mCPR were set lower (42).

District-level covariates were sourced from PSLM 2019-20. Education and income indices followed HDI methodology (43), normalized to 0-1 for comparability. Sectoral composition was measured as the proportion employed in agriculture or manufacturing (services as reference), reflecting the urban-rural gradient and service infrastructure. Fertility was captured as total fertility rate (TFR) among women aged 15-24, indicating early childbearing and unmet need.

Provincial-level random effects were assigned weakly informative half-Student-t priors to allow meaningful between-province variation while preventing overfitting. Correlation between provincial effects for overall CPR and modern CPR was constrained to be near zero, ensuring that traditional and modern contraceptive patterns remain statistically distinct.

Measurement error in administrative proxies was modeled explicitly through province-specific variance components, reflecting bounded but heterogeneous reporting quality across regions. The proxy–outcome relationship was regularized around proportionality, allowing the data to determine deviations while preventing extreme elasticity estimates. To account for differential data quality, the model permitted higher measurement error in predominantly rural districts and lower error in more industrialized districts, consistent with observed differences in reporting infrastructure. Details are presented in Appendix S1.

### 2.4 Posterior Inference and Uncertainty Quantification

The model produced estimates for 121 districts total: 94 districts had crude CPR and modern CPR estimates from Stage 1 that were refined through the hierarchical Bayesian model, while 27 districts lacked crude estimates and were predicted entirely from covariates, provincial random effects, and provincial anchoring constraints. Districts without crude estimates were primarily those with missing, incomplete, or implausible cLMIS data that produced extreme deviations. These included Duki, Harnai, Sohbatpur, Washuk, Quetta, and Sherani in Balochistan; Kohat and Haripur in KP; and several districts in Punjab with data gaps (18 in total), such as Sialkot and Rahim Yar Khan.

For these twenty-seven districts, posterior estimates relied entirely on socioeconomic covariates (education index, income index, fertility rate, sectoral composition), provincial random effects, and provincial anchoring constraints. Uncertainty was quantified using 95% credible intervals derived from the posterior distributions. The width and geographic patterns of these intervals reflect both the quality of input data and the degree of shrinkage applied by the hierarchical model.

### 2.5 Internal and External Validation Checks

Four internal validation approaches assessed model adequacy: (1) Demographic coherence: Provincial rankings matched expected patterns (Punjab > Sindh > KP > Balochistan); (2) Sectoral validation: Confirmed negative agriculture-CPR correlation and positive manufacturing-CPR correlation; (3) Proxy model fit: Correlation between observed and replicated cLMIS data assessed (r>0.7 indicating good fit); (4) Constraint effectiveness: Monitored proportion of districts where modern CPR exceeded overall CPR.

Two external validation approaches assessed estimate statistically coherent: (1) Provincial consistency-district estimates were aggregated to provincial level using MWRA weights and compared to PSLM provincial benchmarks (agreement within ±2 percentage points considered adequate); (2) District-specific validation-model estimates were compared against published district-level CPR values from independent surveys identified through literature review (absolute deviation < 5 percentage points considered consistent).

The administrative boundary shapefiles for Pakistan used in this study were obtained from the geoBoundaries Global Database, an open-access resource provided by the William & Mary geoLab (44).

## Results

The estimated district-level CPR for 121 ranges from 8.6 to 46% and mCPR from 6 to 35%, depicting pronounced variation between and within provinces. Provincial aggregates closely align with PSLM benchmarks (≤ ±0.6 percentage points difference). Validation against the district specific surveys showed that, for 8 out of 12 districts, the mean absolute deviation was under 5 percentage points.

### Model Convergence, Diagnostic Checks and Estimates Variations

Convergence diagnostics indicated that in the hierarchical Bayesian model, the maximum *R* value was 1.003, well below the 1.01 threshold, and both bulk (ESS = 1,456) and tail (ESS = 1,263) effective sample sizes were high, demonstrating strong posterior precision. No divergent transitions or maximum tree-depth saturations were observed, and the mean acceptance statistic (0.95) remained within the optimal range for Hamiltonian Monte Carlo sampling. Bayesian Fraction of Missing Information (BFMI) values for all chains exceeded 0.90, confirming efficient exploration of the posterior energy landscape. Collectively, these diagnostics show that the model converged robustly and that the posterior estimates are stable and reliable (S1 Appendix).

cLMIS-based crude estimates that can have wide dispersion and numerous extreme outliers, reflecting reporting noise and administrative inconsistencies in the raw consumption data. By contrast, Bayesian estimates are smoother, more compact, and more consistent within each province, indicating effective borrowing of strength and correction of measurement error (Figure 2). cLMIS-based crude estimates cLMIS-based crude estimates.

**Figure 2:**
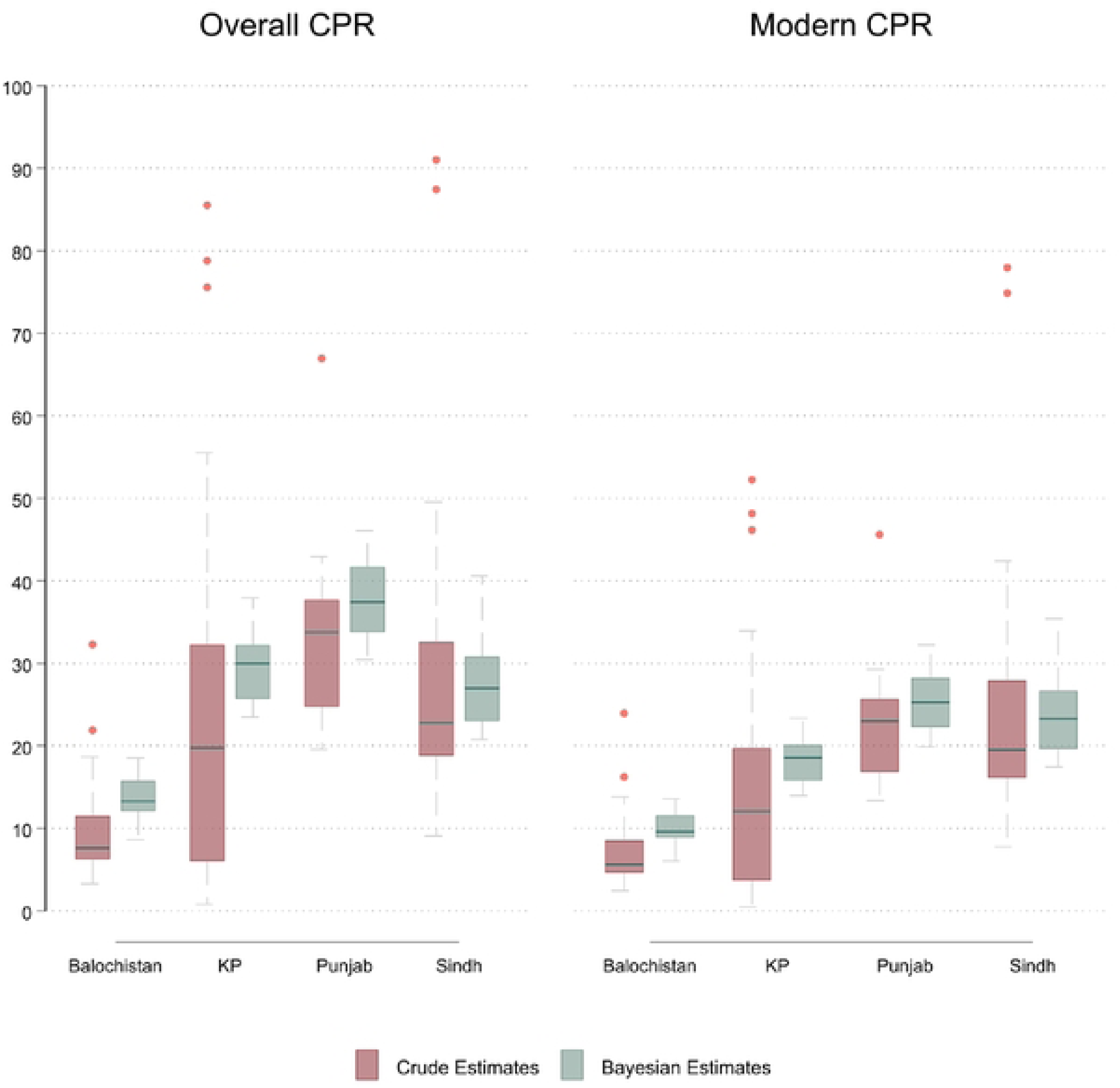
Crude and Bayesian Estimates for CPR and mCPR by Province.

### National and Provincial Validation of Model Outputs

National and provincial estimates were developed by reconverting the total number of users in each district, adding these up and then dividing by the total MWRA population for that province. Table 2 shows that these estimated CPR and mCPR are highly consistent with PSLM survey estimates at both national and provincial levels. National CPR and mCPR differed from PSLM benchmarks by only 0.1 and 0.2 percentage points respectively, and provincial estimates remained within 0.6 percentage points.

**Table 2:** Estimated CPR and mCPR against PSLM Benchmarks.

| Province | Estimated CPR | PSLM CPR | CPR Difference (percentage points) | Estimated mCPR | PSLM mCPR | mCPR Difference (percentage points) |
| --- | --- | --- | --- | --- | --- | --- |
| **National** | 33.8% | 33.7% | +0.1 percentage points | 24.0% | 23.8% | +0.2 percentage points |
| **KP** | 30.0% | 30.6% | −0.6 percentage points | 18.8% | 18.7% | +0.1 percentage points |
| **Punjab** | 38.3% | 38.6% | −0.3 percentage points | 26.0% | 26.3% | −0.3 percentage points |
| **Sindh** | 30.0% | 29.5% | +0.5 percentage points | 25.9% | 25.3% | +0.6 percentage points |
| **Balochistan** | 14.1% | 13.7% | +0.4 percentage points | 10.4% | 10.2% | +0.2 percentage points |

### Secondary External Validation of District Estimates

A secondary validation was attempted, by comparing districts level estimates against available research studies from those localities (Table 3). Across 14 districts where such studies are available, the mean absolute deviation was approximately 4 percentage points. Wider dispersion between these studies may be due to the years since the study was conducted, the limited catchment of the study population and other methodological details. No district-level references for mCPR were identified in the literature, so external validation was limited to overall CPR. Collectively, these findings confirm that the SAE methodology produces credible and externally consistent district-level CPR estimates.

**Table 3:** Comparison of Modelled District CPR Estimates with Literature Values.

| District | Estimated CPR (%) | Literature CPR (%) | Absolute Difference (percentage points) | Study | Survey Year | Validation |
| --- | --- | --- | --- | --- | --- | --- |
| Umer Kot | 28.2 | 25.3 | 2.9 | Muhammad et al., 2025 (45) | 2023 | Adequate |
| Mardan | 31.2 | 40.0 | **8.8** | Nisar et al., 2020 (46) | 2015 | Outside threshold * |
| Bhakkar | 36.9 | 31.9 | 5.0 | Azmat et al., 2015 (47) | 2015 | Borderline adequate |
| Chakwal | 43.8 | 40.6 | 3.2 | Azmat et al., 2015 (47) | 2015 | Adequate |
| Islamabad | 45.3 | 49.0 | 3.7 | Urooj et al., 2022 (48) | 2018 | Adequate |
| Khushab | 38.5 | 28.4 | **10.1** | Tabassum et al., 2016 (49) | 2015 | Outside threshold * |
| Mianwali | 36.3 | 36.6 | 0.3 | Azmat et al., 2015(47) | 2015 | Adequate |
| Okara | 36.2 | 37.4 | 1.2 | Leghari et al., 2022 (50) | 2022 | Adequate |
| Sialkot | 45.7 | 34.0 | **11.7** | Butt et al., 2023 (51) | 2023 | Outside threshold * |
| Badin | 23.0 | 26.1 | 3.1 | Memon et al., 2024 (52) | 2020 | Adequate |
| Matiari | 29.1 | 26.1 | 3.0 | Memon et al., 2024 (52) | 2020 | Adequate |
| Quetta | 17.2 | 33.8 | **16.6** | Sagheer et al., 2019 (53) | 2017 | Outside threshold * |
| **References from old literature** |  |  |  |  |  |  |
| Kohat | 32.2 | 30.8 | 1.4 | Jabeen et al, 2011 (54) | 2011 | Adequate |
| Jamshoro | 26.3 | 29.0 | 2.7 | Bibi et al., 2008 (55) | 2003 | Adequate |
| Tando Allahyar | 25.7 | 29.0 | 3.3 | Bibi et al., 2008 (55) | 2003 | Adequate |

### Uncertainty Assessment

The 95% confidence intervals (CIs) for the district-level estimates indicate moderate and geographically patterned uncertainty. Estimates with CI are presented in S2 Appendix. Across Pakistan, CPR CI widths ranged from approximately 3 to 8 percentage points, while mCPR intervals ranged from about 2 to 7 percentage points. Districts from central Punjab (e.g., Lahore, Faisalabad, Gujranwala) and urban Sindh (e.g., Central Karachi, East Karachi, Hyderabad) that have stronger and more complete cLMIS reporting, exhibited narrower intervals, typically 3-5 percentage points, reflecting more stable information inputs.

Wider confidence intervals were observed in districts with sparser populations, difficult terrain or weak administrative reporting. This applies to districts from Balochistan, including Sherani, Awaran, and Dera Bugti, and the merged districts in KP (formerly the Federally Administered Tribal Area, including Torghar, Mohmand, North Waziristan) that had CPR CI widths closer to 7-8 percentage points. Central and northern districts from Sindh such as Thatta, Sujawal, and Jacobabad also exhibited wider intervals due to known service delivery and reporting limitations.

Overall, the uncertainty levels remained within reasonable bounds, and the hierarchical Bayesian model effectively smoothed extreme fluctuations from Stage 1 while appropriately widening intervals in data-scarce settings. The resulting CI structure aligns with expected patterns of administrative data quality, reinforcing the reliability of the district-level CPR and mCPR estimates.

### District Estimates

The posterior (an updated estimate of a parameter’s value after accounting for observed data as part of a Bayesian analysis) district-level estimates showed substantial variation in contraceptive use across Pakistan. CPR ranged from 8.6% (Dera Bugti) to 46.0% (Rawalpindi and Jhelum), with a national median of 28.9% and an interquartile range of 22.6% to 36.0%. Modern contraceptive use ranged from 6.0% (Dera Bugti) to 35.4% (Central Karachi), with a median of 20.2%.

Figures 3 and 4 show clear spatial disparities in CPR and mCPR across Pakistan. North-Eastern Punjab has the most high-performing districts, with Rawalpindi, Jhelum, Chakwal, Gujrat, Gujranwala, Faisalabad, and Lahore all registering CPR levels above 40%. In contrast, southern Punjab, including Rajanpur, Muzaffargarh, Rahim Yar Khan, and Bahawalpur, shows noticeably lower contraceptive use. Sindh exhibits strong rural-urban contrasts. Karachi districts: Central, East, Korangi, and South Karachi, are among the highest in CPR and mCPR nationwide, while interior Sindh districts such as Thatta, Tando Muhammad Khan, Sujawal, Jacobabad, and Kashmore fall well below the national median.

**Figure 3:**
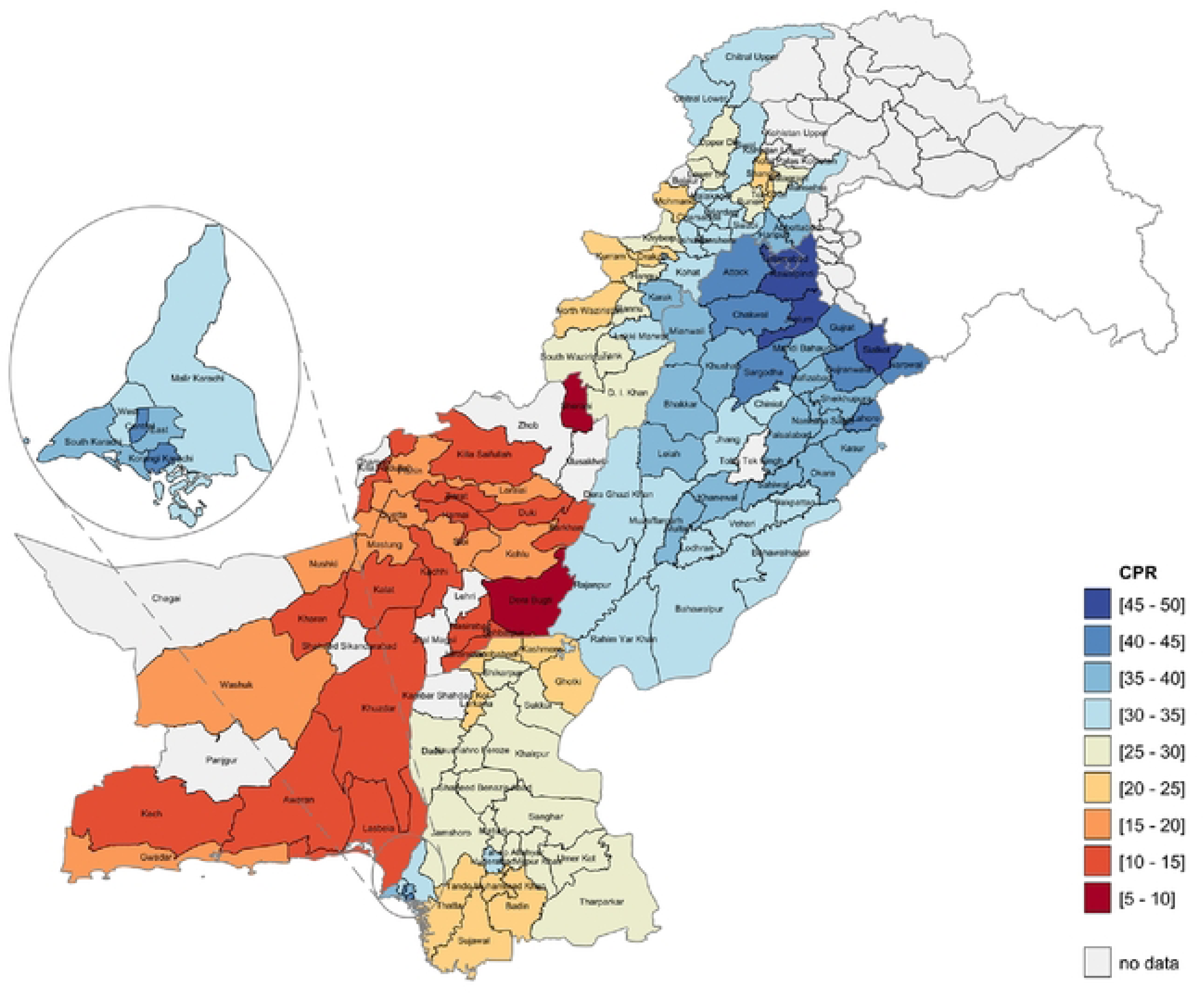
District CPR Across Pakistan. Note: Map Shape Files Source, reprinted from geoBoundaries under a CC BY license, some modifications were made (match recent administrative boundaries) and used for illustrative purposes only.

**Figure 4:**
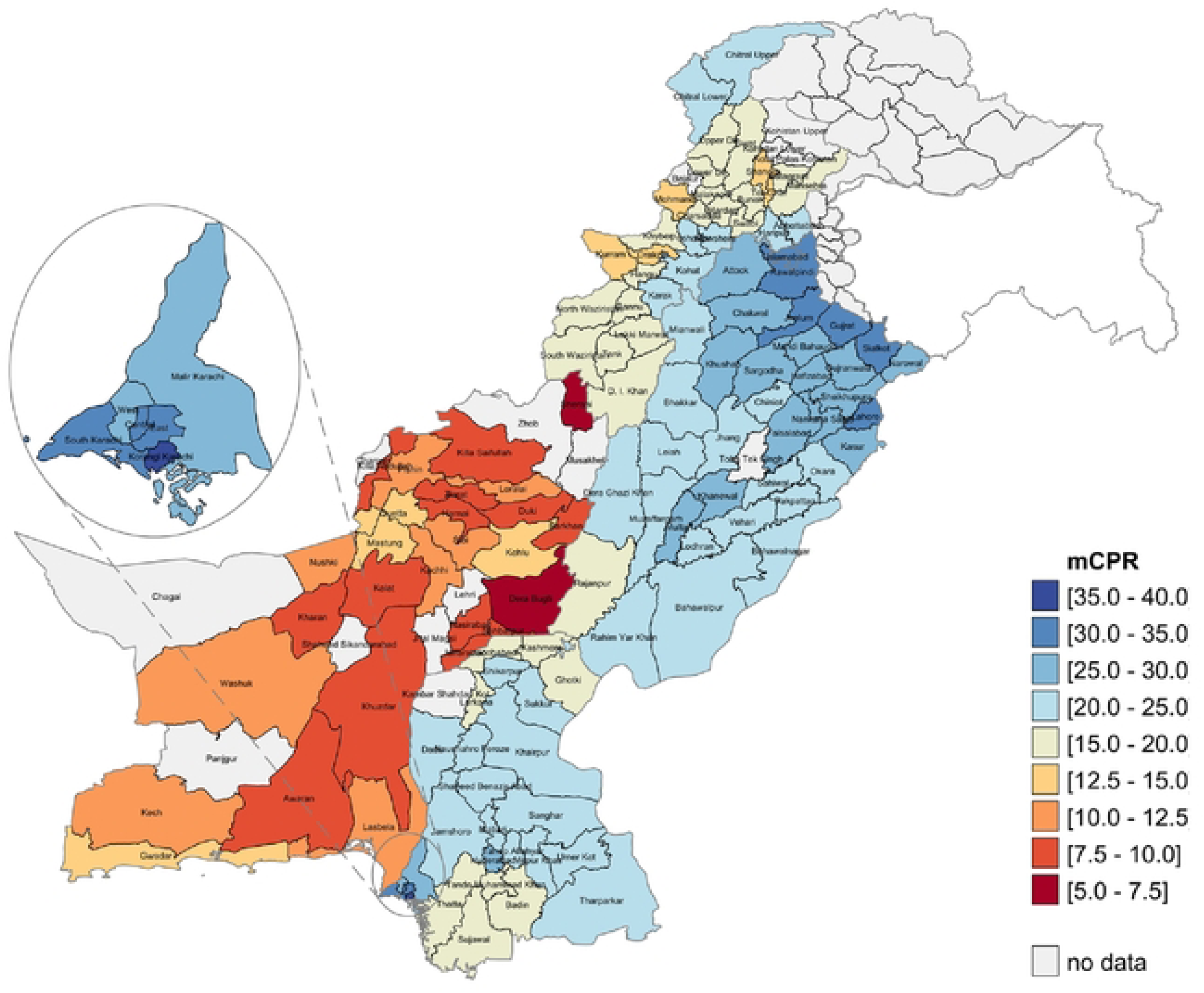
District mCPR Across Pakistan. Note: Map Shape Files Source, reprinted from geoBoundaries under a CC BY license, some modifications were made (match recent administrative boundaries) and used for illustrative purposes only.

In Khyber Pakhtunkhwa, higher CPR and mCPR are seen in the Hazara belt (e.g., Abbottabad, Haripur, and Mansehra), whereas lower uptake is concentrated in mountainous and merged districts (those districts located with KP but were considered “Federally Administered Tribal Areas”, these have recently been merged under KP provincial government, and represent the least developed districts in the province), including Tor Ghar, Shangla, Mohmand, and North Waziristan. Balochistan records the most uniformly low contraceptive use. Districts such as Awaran, Barkhan, Khuzdar, Killa Abdullah, Killa Saifullah, and Sherani exhibit the lowest CPR, with relatively better but still modest values in Gwadar, Kohlu, Mastung, and Quetta.

## Discussion

This study demonstrates that routine logistics data, when formally calibrated within a hierarchical Bayesian small area estimation (SAE) framework, can generate statistically coherent district-level estimates of contraceptive prevalence in Pakistan. Rather than relying solely on survey smoothing, our approach treats administrative supply data as a structured but noisy signal and explicitly models its measurement error. The close alignment of aggregated district estimates with PSLM provincial benchmarks indicates internal coherence, while the stability of posterior intervals suggests that uncertainty has been appropriately quantified rather than suppressed.

In settings where subnational survey data are unavailable, this framework offers a principled alternative for producing policy-relevant local indicators (56). The credibility of the estimates arises from the triangulation of three independent information sources: survey benchmarks (PSLM), demographic denominators (Census MWRA), and district-level consumption signals (cLMIS). Provincial anchoring constrains aggregate drift, while district-level variation is informed by both proxy intensity and socioeconomic covariates. This layered structure ensures that no single data source dominates inference. Districts with weak administrative reporting are partially pooled toward provincial and covariate-informed expectations, whereas data-rich districts retain greater local specificity. This balance between borrowing strength and preserving heterogeneity is central to the robustness of the SAE framework.

The magnitude of district-level variation—exceeding thirty-seven percentage points between the highest- and lowest-performing districts—highlights that contraceptive use in Pakistan is structurally uneven rather than randomly distributed. The observed clustering of higher CPR in economically diversified and urbanized districts (e.g., northeastern Punjab and Karachi) aligns with established associations between female education, household income, service density, and contraceptive uptake. Conversely, persistently low prevalence in agrarian, geographically remote, or infrastructure-constrained districts reflects systemic supply, demand, and sociocultural barriers. Importantly, the spatial pattern suggests that provincial averages mask localized disadvantage, potentially leading to inefficient resource allocation if planning remains provincially aggregated.

From a policy perspective, district-resolution estimates enable more precise targeting of family planning interventions. For example, districts falling below the national median but with moderate socioeconomic capacity may benefit from service-delivery optimization, whereas structurally disadvantaged districts may require broader social and infrastructure investments alongside contraceptive supply expansion (12, 57). The differences are consistent with higher poverty levels, lower education, high pro-fertility preferences and weaker health infrastructure including access to family planning in the latter districts (57–65). Because the model can be updated with routine administrative data, it provides a pathway toward dynamic monitoring rather than episodic survey-based assessment, potentially shifting reproductive health management from reactive to anticipatory planning.

This study extends prior SAE applications in reproductive health in two key ways. First, rather than using administrative data as secondary covariates, we formally treat logistics data as a primary district-level signal while modeling its measurement error through a heteroskedastic proxy structure. Second, the joint estimation of overall and modern CPR introduces cross-indicator borrowing of information, improving stability in data-sparse districts while maintaining conceptual separation between traditional and modern method patterns. This dual-indicator modeling approach enhances inferential efficiency compared with independent estimation strategies. In spatial health applications, SAE has been used to incorporate structured geographic dependence using disease-mapping style models (e.g., BYM and modern re-parameterizations) to improve stability and interpretability of fine-area rates (66, 67). Their use in family planning is more survey-centric, with Bayesian hierarchical models to develop to produce subnational CPR/mCPR patterns and trends, including some incorporation of service statistics (68, 69).

### Limitation

This analysis has several limitations. First, the cLMIS system captures only supply-side data, with incomplete reporting of private-sector provision and no coverage of traditional methods, which may introduce measurement noise despite the model’s adjustment for reporting uncertainty. Second, the estimates rely on 2023 Census MWRA counts as denominators, and any local misreporting can affect precision. Third, Islamabad is grouped within Punjab in the PSLM provincial frame, meaning its district estimates are anchored to Punjab’s benchmark rather than treated as a distinct administrative unit; as a result, its CPR and mCPR values may be slightly over- or under-estimated. Finally, in districts with limited administrative completeness, particularly in Balochistan, rural Sindh, and the merged districts of KP, the model draws more heavily on covariates and provincial patterns, potentially smoothing localized variation. Despite these limitations, the integrated modelling strategy substantially improves upon single-source estimates and provides the most coherent district-level picture possible given the available data systems.

Additionally, provincial anchoring implies that district estimates inherit any systematic bias present in PSLM provincial estimates. While this preserves national coherence, it limits the ability to detect province-wide underestimation or overestimation. Future work incorporating independent validation surveys or alternative national benchmarks could further strengthen calibration.

## Conclusion

In summary, this study demonstrates that Bayesian small area estimation can convert fragmented administrative and survey data into coherent district-level reproductive health indicators. The approach reveals substantial geographic inequities that are obscured by provincial averages and provides a statistically principled framework for routine subnational monitoring in data-constrained settings. While not a substitute for high-quality household surveys, this integrated modeling strategy represents a pragmatic complement, enhancing the resolution, frequency, and policy relevance of contraceptive prevalence measurement in Pakistan and comparable LMIC contexts.

## Data Availability

The data underlying the results presented in the study are drawn from third-party sources. PSLM and Census data are owned by the Pakistan Bureau of Statistics and are available upon request from https://www.pbs.gov.pk. cLMIS administrative data are owned by government health authorities and cannot be shared publicly due to data governance and confidentiality restrictions. Aggregated outputs and analytical results are provided within the manuscript and Supporting Information files.

## Acknowledgment

We would like to extend our gratitude to CHEMONICS Pakistan for providing the valuable data used in this study. Their contribution has been instrumental in enabling our research and enhancing our understanding of the supply and dispensation dynamics of contraceptives in Pakistan.

---

**S1 Appendix: Methodology, Model Convergence and Diagnostics**

**S2 Appendix: District Estimates of CPR and mCPR across Pakistan**

